# Anorexia nervosa polygenic risk, beyond diagnoses: relationship with adolescent disordered eating and behaviours in an Australian female twin population

**DOI:** 10.1101/2023.11.06.23298183

**Authors:** Madeleine Curtis, Lucia Colodro-Conde, Sarah E. Medland, Scott Gordon, Nicholas G. Martin, Tracey D. Wade, Sarah Cohen-Woods

## Abstract

**Background:** It is well established that there is a substantial genetic component to eating disorders. Polygenic risk scores (PRS) can be used to quantify cumulative genetic risk for a trait at an individual level. Recent studies suggest PRS for Anorexia Nervosa (AN) may also predict risk for other disordered eating behaviours, but no study has examined if PRS for AN can predict disordered eating as a global continuous measure. This study aimed to investigate whether PRS for AN predicted overall levels of disordered eating, or specific lifetime disordered eating behaviours, in an Australian adolescent female population.

**Methods:** PRS were calculated based on summary statistics from the largest Psychiatric Genomics Consortium AN genome-wide association study to date. Analyses were performed using GCTA to test the associations between AN PRS and disordered eating global scores, avoidance of eating, objective bulimic episodes, self-induced vomiting, and driven exercise in a sample of Australian adolescent female twins recruited from the Australian Twin Registry (N = 383).

**Results:** After applying the false discovery rate correction, the AN PRS was significantly associated with all disordered eating outcomes.

**Conclusions:** Findings suggest shared genetic aetiology across disordered eating presentations and provide insight into the utility of AN PRS for predicting disordered eating behaviours, and individuals at risk, in the general population. In the future, PRSs for eating disorders may have clinical utility in early disordered eating risk identification, prevention, and intervention.

## Introduction

Disordered eating is a term that encompasses all eating disorders described in the DSM-5, as well as disordered eating-related behaviours and cognitions that may not meet criteria for any eating disorder diagnosis, but still cause significant distress and impairment (American Psychiatric Association [APA], 2013). This includes cognitions such as an intense fear of weight gain or a distortion in body image, and behaviours such as food restriction, binge-eating, or self-induced vomiting (APA, 2013). Both behavioural and cognitive disordered eating symptoms can cause significant physical and psychological impairment, regardless of whether criteria for a specified clinical eating disorder diagnosis is met (APA, 2013; Wilkop, Wade, Keegan, & Cohen-Woods, 2023).

Disordered eating typically develops during adolescence and continues to disproportionately affect females, with a recent study reporting point prevalence of any ED among Australian adolescents of 32.9% for females and 12.8% for males (Mitchison et al., 2020). Eating disorder presentations are dynamic and can change with both age and duration (Forbush et al., 2018; Milos, Spindler, Schnyder, & Fairburn, 2005). For example, Forbush et al. (2018) reported that, in their sample, 100% of those who had an initial diagnosis of anorexia nervosa (AN) or binge-eating disorder (BED), and 50% of those initially diagnosed with bulimia nervosa (BN), met criteria for a different eating disorder diagnosis after one year. A change in diagnosis was also seen in 23.9% of those initially diagnosed with other specified feeding or eating disorder (OSFED), suggesting progression to full criteria eating disorder (Forbush et al., 2018). Milos et al. (2005) similarly reported high rates of crossover between eating disorder diagnoses, with only 48% of participants with AN, 27% of those with BN, and 10% of those with eating disorder not otherwise specified retaining their initial diagnosis over a 30-month follow up period.

There is clear evidence that restrictive eating disorders first emerge in early adolescence, with binge/purge symptoms developing later in adolescence or early adulthood (Fairburn, Cooper, & Shafran, 2003; Hudson, Hiripi, Pope, & Kessler, 2007). This age-based disordered eating trajectory likely reflects changes in eating behaviours, rather than changes in cognitive symptoms which typically remain the same (Fairburn et al., 2003). Disordered eating symptoms often exist prior to the development of a clinical eating disorder, with milder symptoms in childhood worsening through adolescence to adulthood (McClelland, Robinson, Potterton, Mountford, & Schmidt, 2020), again suggesting an age-related trajectory. Consequently, there is a need for research that both considers the broader construct of disordered eating, which is not restricted to specific diagnostic criteria, and captures measurements across adolescence, a dynamic period of growth and change.

Psychiatric disorders are polygenic in nature, meaning that thousands of genetic variants contribute to the risk for developing psychiatric disorders, with each variant having a very small, additive effect (Visscher, Yengo, Cox, & Wray, 2021). Because of this, genome-wide association studies (GWASs) are the preferred method for identifying genetic risk factors for polygenic traits, and involve testing hundreds of thousands of genetic variants that span the entire genome to identify which variants are most associated with the trait of interest (Hübel, Leppä, Breen, & Bulik, 2018). GWASs on eating disorders are currently limited due to the large samples required. To our knowledge, AN is currently the only clinically diagnosed eating disorder to have been the subject of large GWASs, although recruitment for GWASs on BN and BED are underway (Bulik et al., 2022; Steiger & Booij, 2020). A GWAS on BED has been conducted recently, however cases were predicted using machine learning as clinical BED diagnoses were not collected (Burstein et al., 2022). Machine learning was applied to distinguish between clinically diagnosed BED and obesity, but this does mean there may be some misassignment.

The largest GWAS of AN to date was conducted by Watson et al. (2019), who identified 8 genome wide significant loci associated with AN. It was estimated that 11-17% of phenotypic variation could be attributed to single nucleotide polymorphism (SNP) variation captured, however the polygenic risk score (PRS) generated from this GWAS accounted for just 1.7% of phenotypic variation (Watson et al., 2019). This means there are still many more risk variants for AN to identify, which will likely happen as sample sizes increase as this will improve power to identify significant associations (Bulik et al., 2022). The Watson et al. (2019) GWAS did not observe any differences in genetic variation between those with different AN subtype, specifically with and without binge eating. Although these presentations come under the same diagnostic category of AN, this indicates that it is worthwhile to investigate if other eating disorders also share a similar genetic basis to AN. This suggestion is consistent with findings of a twin study showing significant overlap in genetic risk factors between a group with either lifetime AN, BN or BED and a group with OSFED (Fairweather-Schmidt & Wade, 2014). However, no study to date has investigated the relationship between AN PRS and disordered eating more broadly as a phenotype within the population.

This study aimed to investigate whether PRS for AN could predict disordered eating in an Australian female twin population. A small number of studies have investigated whether AN PRSs can predict other specific disordered eating behaviours, however findings have been mixed and restricted to two cohorts. One study investigated the Adolescent Brain and Cognitive Development cohort at ages 9 – 11 and reported no relationship between AN PRS and eating disorder psychopathology, based on a lifetime eating disorder screener (Westwater et al., 2023). Similarly, Abdulkadir and colleagues (2022) reported no association between AN PRS and eating disorder symptoms in the ALSPAC cohort based on the youth risk behaviour surveillance questionnaire at ages 14, 16, and 18. Both studies combined boys and girls. In contrast, Yilmaz and colleagues (2022) also investigated the ALSPAC cohort, but it was slightly larger and included sex-specific analyses. In this study they identified a relationship between AN PRS and EDNOS / purging disorder, presence of any eating disorder, and compulsive exercise, at age 14 in girls specifically.

All these studies categorised individuals with dichotomous yes/no responses in relation to disordered eating behaviours, and also relied on parent report (except for ALSPAC age 16 and 18 measures). No studies have considered disordered eating behaviours as a broad continuous phenotype, using global measures of eating psychopathology. The high levels of impairment experienced by those with disordered eating, even when not meeting diagnostic criteria for a specified eating disorder, means that research on the broader phenotype of disordered eating is important (Wilkop et al., 2023). As eating disorder presentations often change with both age and duration, whereas genetic variants captured by GWASs are stable from birth (Brookes, 1999; Fairburn et al., 2003; Forbush et al., 2018), it will be helpful to ascertain if genetic variants associated with AN are also associated with disordered eating broadly. If PRS can quantify genomic risk for disordered eating at an individual level, then PRS may have high clinical utility for targeted early intervention and prevention efforts in the future. To explore this, we tested whether AN PRS predicted overall levels disordered eating. As there have also been limited studies looking at specific disordered eating behaviours, and to see if any association found between AN PRS and overall disordered eating is driven by specific disordered eating behaviours, we additionally tested whether a higher AN PRS was associated with any lifetime disordered eating behaviours, specifically avoidance of eating, objective bulimic episodes, self-induced vomiting, or driven exercise.

## Method

### Sample

The target cohort for the present study consisted of Australian adolescent female twins. Data were collected over three waves with the data collection process described in detail in previously published studies (Fairweather-Schmidt & Wade, 2014, 2017; Wade, Byrne, & Bryant-Waugh, 2008; Wade et al., 2013; Wade & O’Shea, 2015; Wilksch & Wade, 2009). Ethical approval was provided by the Flinders University Clinical Research Ethics Committee (no. 115/07) and written informed consent was obtained from all participants. Data were collected longitudinally over three time periods (Wave 1, Wave 2, Wave 3), as shown in Figure 1. Participants eligible for inclusion at the time of data collection were female-female twin pairs who were registered with the Australian Twin Registry (ATR) and aged between 12 and 15 years at the first wave of data collection. Wave 1 included 669 participants (mean age 13.96, SD = 0.80), Wave 2 included 669 (mean age 15.1, SD = 0.83), and Wave 3 included 499 participants (mean age 16.9, SD = 0.70). There was a mean duration of 1.5 years (SD = 0.17) between Waves 1 and 2, and 2.96 years (SD = 0.27) between Waves 2 and 3.

**Figure 1.**
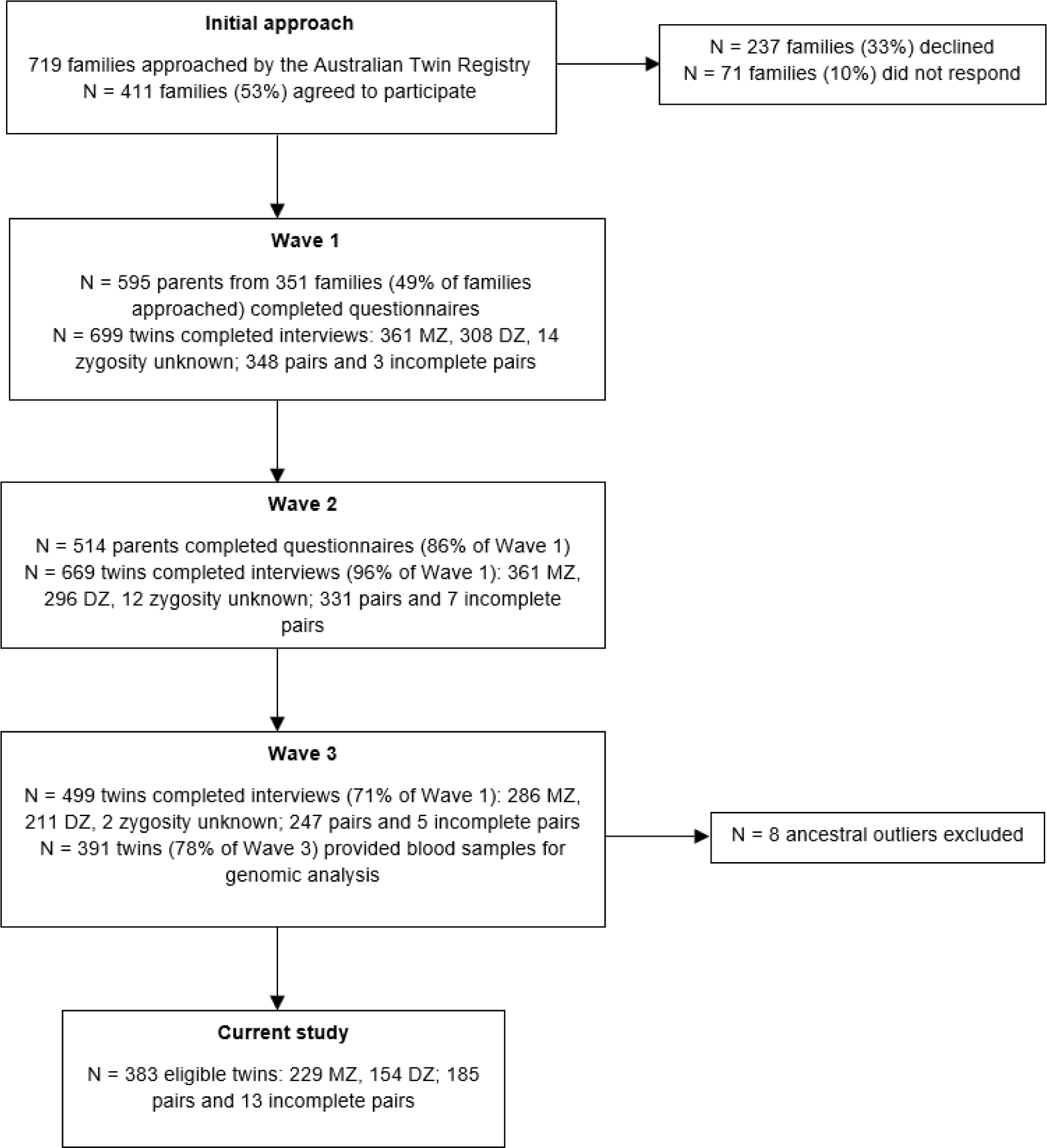
Flow diagram of data collection.

### Procedure

#### Wave 1

A total of 719 families were approached by the ATR. Of these, 411 (57.2%) agreed to participate in the study, 237 (32.9%) declined, and 71 (9.9%) did not respond. Those who agreed to participate were approached by researchers. Both parents of each set of twins were sent self-report questionnaires. Questionnaires were returned by 595 parents (348 mothers and 247 fathers) from 351 families (85.4% of families who agreed to participate). When questionnaires were returned from at least one parent, the twins were administered the Eating Disorder Examination (EDE) verbally via telephone, with each twin in the family interviewed at a separate time and by a different clinician. A total of 699 twins (348 complete pairs and 3 incomplete pairs) participated and completed telephone interviews at Wave 1.

#### Wave 2

All eligible twins were contacted by the ATR and invited to participate in Wave 2, including non-responders from Wave 1. A total of 669 twins agreed to participate (96% of Wave 1). Wave 2 used the same procedure for data collection as Wave 1, including administering the self-report questionnaire to parents and conducting the telephone interviews with twins.

#### Wave 3

All eligible twins were contacted by the ATR and invited to participate, including non-responders from Waves 1 and 2. A total of 499 twins (71% of Wave 1) agreed to participate. The telephone interview was administered to all twins and blood samples were provided for analysis by 391 twins (78% of Wave 3).

Eight participants were excluded due to being ancestral outliers, leaving a total of 383 eligible participants. The included sample consisted of 185 twin pairs and 13 incomplete twin pairs.

### Disordered Eating Measures

Participants’ global scores on the 12^th^ edition of the Eating Disorder Examination (EDE) (Fairburn, Cooper, & O’Connor, 1993) were used as a continuous measure of disordered eating. The EDE was slightly modified for use with children in line with previous recommendations (Bryant-Waugh, Cooper, Taylor, & Lask, 1996; Wade et al., 2008). The global score includes four subscales (Eating Concern, Weight Concern, Shape Concern, Dietary Restraint) and measures disordered eating over the previous 28 days. The EDE was administered to participants as a semi-structured interview via telephone at each wave of data collection. The global score consisted of 22 items measured on a 7-point Likert scale (scores ranging 0-6), calculated as the mean score across all items. Global EDE scores are a useful and widely utilised measure of eating pathology, and studies have found the global scores to be a stronger indicator of overall eating pathology than individual subscale scores (Friborg, Reas, Rosenvinge, & Rø, 2013; Jenkins & Rienecke, 2022). Global EDE scores have also been shown to accurately discriminate between those with and without a clinically diagnosed eating disorder (Aardoom, Dingemans, Slof Op’t Landt, & Van Furth, 2012; Mond et al., 2008). To ensure we captured the greatest amount of eating pathology experienced by each participant, we used the highest global EDE score recorded across all 3 waves of data collection as our continuous measure of disordered eating.

The EDE telephone interview also included several behavioural frequency questions, which have demonstrated high interrater reliability across studies (Berg, Peterson, Frazier, & Crow, 2012). The behavioural frequency questions addressed a three-month period and assessed the presence of both current and lifetime disordered eating behaviours. As we were interested in disordered eating behaviours at any point throughout the participants’ life, we used the lifetime diagnostic questions. We considered the lifetime disordered eating behaviours to be present (yes/no) if there was ever a time that the participant had engaged in the disordered eating behaviour for a three-month period. Four lifetime behaviours were investigated: avoidance of eating, objective bulimic episodes, self-induced vomiting, and driven exercise. Three studies investigating AN PRS have used the presence of these behaviours as disordered eating outcomes (Abdulkadir et al., 2022; Westwater et al., 2023; Yilmaz et al., 2022). However, no study has investigated whether AN PRS predicts the lifetime occurrence of these disordered eating behaviours in a female cohort.

### Data Analysis

DNA samples from the target sample were genotyped at Erasmus Medical Centre (Rotterdam) using the Infinium Global Screening Array V.1 (Illumina, CA). Raw genotypes were called using standard procedure in GenomeStudio as part of a large batch of 10,656 samples. These were exported on the Illumina ‘TOP’ strand and checked for samples with low call rate (below 97%) and identifiable sex errors and IBD state with respect to the twins (no issues identified for these samples). Markers were filtered to remove a set of 560 markers which were duplicates or non-alignable to hg19, then those with GenTrain score < 0.6; missingness >5%; MAF < 1%; low Hardy-Weinberg p-value (<10^-6^) or failing sex-chromosome-specific filters (chrX: heterozygosity >1% for males), removing 189,039 markers; keeping 502,739. This batch was integrated with other identically quality-controlled batches from GSA chips, converted to the hg19 plus strand, and imputed for the 432,255 markers passing QC in all batches, using default settings and the HRCr1.1 reference on the Michigan Imputation Server (phased using Eagle; imputed using minimac4).

The PRSs for AN were calculated in PLINK version 1.9 (Chang et al., 2015) using GWAS summary statistics obtained from the Eating Disorders Working Group of the Psychiatric Genomics Consortium (PCG-ED). We used the Freeze 2 AN sample, which combined 33 cohorts to include data from 16,992 AN cases and 55,525 controls (Watson et al., 2019). Australian and New Zealander participants (N = 2,536 AN cases and 15,624 controls) were excluded from our PRS calculations to avoid potential crossover between base and target datasets. We performed clumping to account for linkage disequilibrium (LD) among SNPs and extract genetically independent variants. We retained the SNP with the smallest *p* value in each 1000kb window and removed those in LD with this SNP (*r^2^* > 0.1). PRS were calculated using the sum of all retained SNPs weighted by their effect sizes as estimated in the GWAS summary statistics (Watson et al., 2019). PRS were calculated for eight different *p* value thresholds (*p* < 5 x 10^-8^, *p* < 1 x 10^-5^, *p* < 0.001, *p* < 0.01, *p* < 0.05, *p* < 0.1, *p* < 0.5, *p* < 1.0). Genome-wide complex trait analysis (GCTA) genome-based restricted maximum likelihood (GREML) analyses (Yang, Lee, Goddard, & Visscher, 2011) were used to test the association between AN PRS and global disordered eating scores, and the presence of lifetime disordered eating behaviours, at the eight *p* value thresholds, controlling for covariates. We controlled for relatedness using GCTA to calculate the genetic relationship matrix (GRM) for our target sample. The first five genetic principal components (PCs) were included as covariates to control for population stratification (Price et al., 2006). Additional covariates included were participant age and BMI centile. Age was included to control for age-related differences in disordered eating (Fairburn et al., 2003; McClelland et al., 2020) and BMI has been associated with disordered eating behaviours and attitudes (Goldschmidt, Aspen, Sinton, Tanofsky-Kraff, & Wilfley, 2008). We used BMI centile (BMI-for-age) in place of BMI as it is considered more accurate than BMI for children (CDC, 2022). We corrected for multiple testing by calculating the false discovery rate (FDR) adjusted *p* values (*Q*) using the Benjamini and Hochberg method (Benjamini & Hochberg, 1995).

## Results

### Participant Characteristics

Participant characteristics are displayed in Table 1. The mean age of the included participants was 14.01 (*SD* = 0.78) at Wave 1, 15.15 (*SD* = 0.82) at Wave 2, and 16.95 (*SD* = 0.83) at Wave 3, with an overall range of 12.74 – 19.84 years. Participants were of European ancestry and had an average Socioeconomic Indexes for Areas (SEIFA) of 100.95 (*SD* = 10.80). SES was calculated using information on parental income, education, and occupation to produce a standardised measure of socioeconomic status (*M* = 100, *SD* = 15) (Farish, 2004).

**Table 1.**
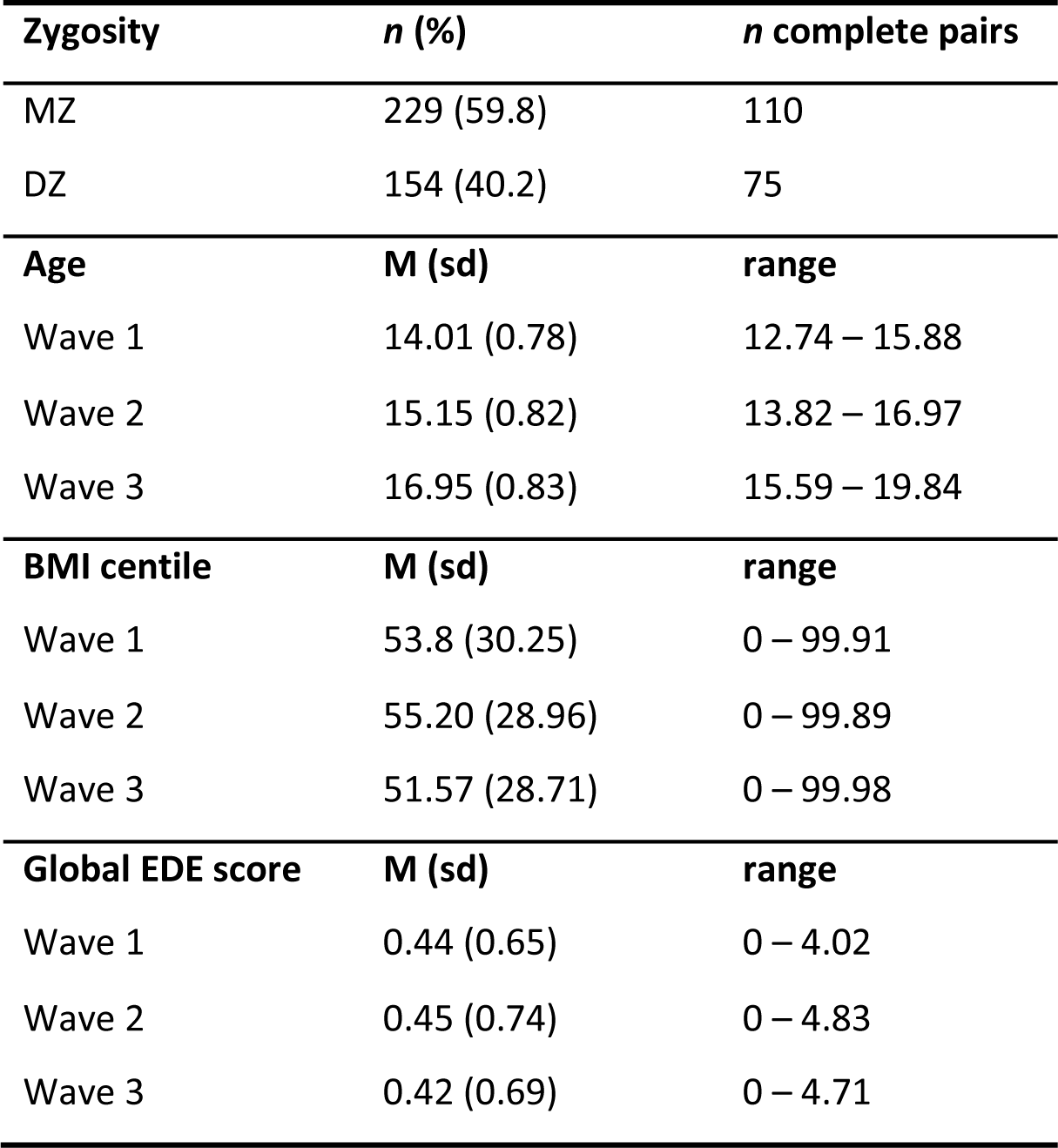
Participant characteristics.

### Disordered eating global score

The results of all PRS association analyses are presented in Figure 2. After applying the FDR correction, the AN PRS significantly predicted global disordered eating in the target sample at five of the eight *p* value thresholds, with the most significant association at the *p* < 0.5 threshold (*r^2^* = 2.19%, *Q* = 0.015).

**Figure 2.**
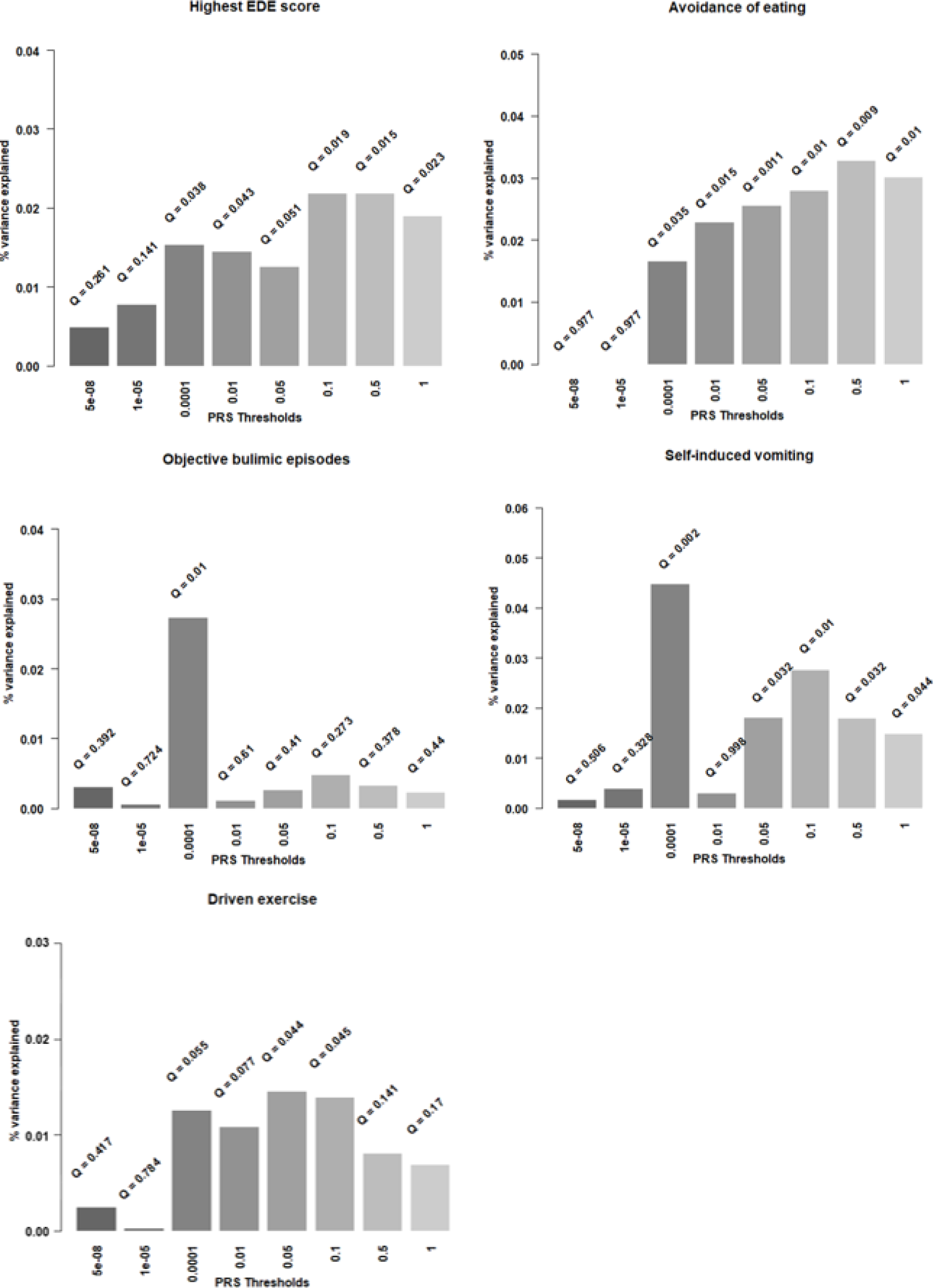
Associations between AN PRS and disordered eating outcomes at eight p value thresholds.

### Lifetime disordered eating behaviours

The lifetime prevalence in our target sample for avoidance of eating, objective bulimic episodes, self-induced vomiting, and excessive exercise were 5%, 4.2%, 1.8% and 7.8% respectively. Avoidance of eating was significantly associated with the AN PRS at six *p* value thresholds, with *p* < 0.5 generating the most significant association (*r^2^* = 3.29%, *Q* = 0.009). The AN PRS significantly predicted both self-induced vomiting (*r^2^*= 4.48%, *Q* = 0.002) and objective bulimic episodes (*r^2^*= 2.74%, *Q* = 0.010) at *p* < 0.001. Excessive exercise was also significantly associated with the AN PRS after applying the FDR correction at both *p* <0.05 (*r^2^* = 1.46%, *Q* = 0.044) and *p* < 0.1 (*r^2^* = 1.40%, *Q* = 0.045).

## Discussion

This study was the first to comprehensively explore whether AN PRS predicts disordered eating as a broad phenotype in the general population. This was achieved by investigating both global continuous measures of disordered eating, as well as four specific lifetime disordered eating behaviours. This approach enabled us to examine whether any association between AN PRS and disordered eating was specific to restrictive behaviours or extended to disordered eating behaviours of relevance across different eating disorders. After applying the false discovery rate correction, the AN PRS was still associated with all disordered eating outcome measures, explaining a significant amount of phenotypic variance in global EDE scores (2.19%), avoidance of eating (3.29%), objective bulimic episodes (2.74%), self-induced vomiting (4.48%), and driven exercise (1.46%). Our finding that the AN PRS predicted global disordered eating scores in our sample supports the possibility of a shared genetic basis across different eating disorder presentations. Given that this is the first study to apply the AN PRS to a global disordered eating measure, this is a novel finding that is worthy of further exploration in larger and more diverse target cohorts. It is notable that the phenotypic variance captured by the AN PRS in global EDE scores in this study is equivalent to the phenotypic variance reported by Watson et al (2019) for clinical AN cases. Further, phenotypic variance for feeding-related behaviours captured by the AN PRS was even greater. This may be attributable to the broader inclusion of individuals who may be classified unaffected in conventional case-control studies as they have not received a diagnosis of AN (or other eating disorder), but who may sit very close to the boundary of that classification. This highlights the importance of investigating broader population-based phenotypes such as disordered eating, which includes individuals that would otherwise be misclassified as unaffected or potentially excluded from eating disorder studies.

To our knowledge, there are currently no published studies that explore the association between AN PRS and a global measure of disordered eating, and limited studies that use AN PRS as a predictor for specific disordered eating behaviours. Existing studies have investigated avoidance of eating, self-induced vomiting, and excessive exercise behaviours, but the present study is the first to additionally include objective bulimic episodes as an outcome. Consistent with our results, Yilmaz et al. (2022) reported a significant association between the AN PRS and excessive exercise. However, in a study by Abdulkadir et al. (2022) using the same target cohort from the Avon Longitudinal Study of Parents and Children (ALSPAC) as Yilmaz et al. (2022), a significant association was not reported. As the target sample and measures were the same across the two studies, the difference in results is likely explained by sex differences. Abdulkadir et al. (2022) used a combined male and female ALSPAC cohort, whereas Yilmaz et al. (2022) investigated the predictive ability of the AN PRS in males and females from the ALSPAC cohort separately, with the AN PRS significantly predicting compulsive exercise in females, but not in males. As the present study also used a female cohort, this suggests that driven exercise may be a phenotype that has a higher genetic component in females compared to males.

Our finding that AN PRS predicted avoidance of eating and self-induced vomiting behaviours contrasts with the studies by Abdulkadir et al. (2022), Yilmaz et al., (2022) and Westwater et al. (2023), who all reported no significant associations between AN PRS and these behaviours. Although these studies used the same GWAS summary statistics (Watson et al., 2019) to generate the AN PRS scores as our study, there were differences in the measures of disordered eating behaviours across studies that may have contributed to our contrasting findings. Both Yilmaz et al. (2022) and Abdulkadir et al. (2022) used self-report measures of symptoms that occurred over the previous year, whereas our study utilised a clinician-administered interview and assessed the lifetime presence of disordered eating behaviours. Westwater et al. (2023) also used lifetime endorsement of disordered eating behaviours, however this was assessed through parent ratings using screener items, reducing reliability. This likely increased our power to detect significant findings in comparison to the other studies, despite our much smaller target sample size.

The discrepancies in our findings compared to Westwater et al. (2023) may also partially be explained by participant age differences. Participants were younger (age 9-11 years) in the study by Westwater et al. (2023), compared to the present study (age 12-19 years). Disordered eating behaviours typically emerge during adolescence, with bulimic-type behaviours such as self-induced vomiting often developing in early adulthood (Fairburn et al., 2003). Twin studies have also identified differences in genetic contributions to disordered eating across adolescence, with genetic influences increasing with age (Fairweather-Schmidt & Wade, 2015; Klump, Burt, McGue, & Iacono, 2007; Klump, McGue, & Iacono, 2000; O’Connor, Culbert, Mayhall, Burt, & Klump, 2020). This means the present study is likely to have captured both an increase in disordered eating behaviours and an increase in genetic contribution among participants, who were further along in adolescence compared to Westwater et al. (2023).

Limitations need to be considered when interpreting our results. Despite finding significant associations between the AN PRS and all disordered eating outcomes, it is likely that our analyses were still underpowered due to the sample size of the AN GWAS used to generate the PRS (Watson et al. 2019). The AN PRS at this stage is relatively limited in statistical power, with the predictive power of a PRS driven by the sample size of the respective GWAS, meaning larger sample sizes will increase power to detect robust significant associations (Hübel et al., 2018). This has been demonstrated in genomic studies on several psychiatric disorders, with the most notable being schizophrenia (Pantelis et al., 2014; Smoller et al., 2019; Wray et al., 2018). GWAS investigating variants associated with AN have only recently been sufficiently powered to detect any loci significantly associated with AN at the genome-wide level (Watson et al., 2021; Wray et al., 2014). Initial GWAS on AN failed to identify any genome-wide significant loci associated with AN, with the first large-scale GWAS on AN using a cohort of 1,033 AN cases and 3,733 controls (Wang et al., 2011). Duncan et al. (2017) were the first to identify a significant locus with a cohort of 3,496 AN cases and 10,982 controls, and the most recent GWAS by Watson et al. (2019) identified eight genome-wide significant loci, with many additional loci also close to the genome-wide significance threshold. This was achieved by combining 33 datasets, resulting in a total sample of 16,992 cases and 55,525 controls and demonstrating that as AN GWAS sample sizes increase, the power to detect significant associations will also increase (Watson et al., 2021). Of course, the PRS utilises loci that fall below genome-wide significance, but power of PRSs significantly improves as the discovery cohort sample size increases.

Our target sample was relatively small, as it was limited by the number of participants who provided blood samples for analyses and had genomic data available. Replication in a larger adolescent female target cohort using the same global EDE and lifetime behaviour outcome measures will enable more powerful analyses. A strength of the present study was the use of a target sample from the general population, however this also meant there was a low number of participants who displayed disordered eating symptoms, potentially reducing the power of our analyses. The mean global EDE score of our target sample was low (0.44 across all waves) and the number of participants endorsing lifetime behaviours was small, however the prevalence of lifetime behaviours in our sample was in line with norms in comparable populations (Aardoom et al., 2012; Luce, Crowther, & Pole, 2008; Machado et al., 2014; Mond, Hay, Rodgers, & Owen, 2006), and as previously mentioned the phenotypic variance in disordered eating captured by the AN PRS was equivalent to that reported in the most recent large GWAS (Watson et al., 2019).

Finally, it is important to highlight that the present study used a target sample of adolescent females of European ancestry, so findings cannot be generalised to other ages, genders, or ethnicities. This was further limited by availability of AN GWAS data, which is currently restricted to individuals of European ancestry (Watson et al., 2019). Eating disorder research has primarily focused on females, however this is not representative of the diverse group of people who experience disordered eating (Huckins et al., 2022). Future studies need to utilise broader or more diverse recruitment of people with eating disorders so findings can be more representative of those who experience symptoms.

Overall, the present study demonstrates that AN PRS have the ability predict disordered eating behaviours beyond those listed as AN diagnostic criterion, suggesting a shared genetic component across different disordered eating behaviours. This means the AN PRS may be useful in identifying individuals at greater risk of developing any disordered eating symptoms. Whilst it is critical to note that at this time AN PRS does not have utility as a diagnostic or predictive tool, as presently it captures only a small part of the genetic contribution to our phenotypes, in time this will increase. As we work towards improved predictive power, this work contributes to growing literature that supports the use of PRS in identifying individuals at risk for sub-threshold behaviours, and in turn fully developed conditions, based on greater genetic susceptibility. Combining polygenic risk with other environmental and social risk factors in the future has promise to develop useful models where early interventions may be directed.

## Data Availability

All data produced in the present study are available from the corresponding author upon reasonable request.

## Data

All the data produced in the present study are available from the corresponding author upon reasonable request.

## Acknowledgements

We acknowledge and thank all the participants in our study, for their time and willingness to engage with research.

